# Atopy, asthma symptoms, and eosinophilic airway inflammation in British woodworkers

**DOI:** 10.1101/2025.03.06.25321879

**Authors:** Ruth Wiggans, Jade Sumner, Ed Robinson, Charlotte Young, Andrew Simpson, Timothy Yates, David Fishwick, Christopher Michael Barber

## Abstract

**Objectives:** Although wood dust remains a leading cause of occupational asthma (OA) in Great Britain, there have been no recent studies in British woodworkers. This cross-sectional study examined asthma risk factors in woodworkers across exposure groups.

**Methods:** Participants answered a respiratory questionnaire and underwent fractional exhaled nitric oxide (FE_NO_), spirometry, and specific immunoglobulin E measurements. Wood dust exposure was assigned through a specific job-exposure matrix. Multiple regression evaluated asthma risk factors identified a priori including wood dust exposure, atopy, and current asthma symptoms.

**Results:** A total of 269 woodworkers participated. Median wood dust exposure was 2.00mg/m^3^ (IQR 1.14 mg/m^3^). Current asthma symptoms (CAS), work-related respiratory symptoms (WRRS) and eosinophilic airway inflammation (FE_NO_ >40ppb) were common, present in 46%, 11% and 19% of the cohort, respectively. Atopic woodworkers were at increased risk of WRRS (OR 2.78, 95% CI 1.11 – 6.92, p<0.05), asthma (OR 3.40, 1.49 – 7.81, p<0.01), and FE_NO_ >40ppb (unadjusted OR 2.00, 1.03 – 3.88, p<0.05). No effect was seen for airflow obstruction. Workers with CAS were at increased risk of WRRS and ever asthma (4.29, 2.12 – 8.69, p<0.001) but not FE_NO_ >40ppb or airflow obstruction. Increasing exposure did not significantly increase risk of asthma symptoms, asthma, airflow obstruction and inflammation.

**Conclusions:** Asthma symptoms were prevalent among British woodworkers, even at low exposure levels. Atopy significantly increased asthma risk, particularly among symptomatic woodworkers. Further studies to phenotype and endotype populations of workers at risk of, and suffering from, wood dust OA will inform future approaches to screening and diagnosis in these populations.

**What is already known on this topic:** Wood dust is a leading cause of occupational asthma (OA) in Great Britain. No recent studies have described risk factors for OA in British woodworkers. Evidence identifies atopy, asthma symptoms, and wood species as risk factors for OA, but not consistently so.

**What this study adds:** This cross-sectional study used a detailed job-exposure matrix, questionnaire and clinical data to understand risk factors for asthma in British woodworkers. We found upper airway, asthma symptoms (CAS), work-related symptoms and eosinophilic airway inflammation to be common among British woodworkers, but specific sensitisation to wood dust was low. Among workers with asthma symptoms atopy significantly increased the risk of CAS, asthma, and airway inflammation in woodworkers. Increasing wood dust exposure was not associated with an increased risk of asthma symptoms or asthma.

**How this study might affect research, practice or policy:** This research provides the first epidemiological study on asthma in British woodworkers for decades and highlights specific risk factors for asthma in this group. This data is useful to inform health surveillance programmes and may help inform any future review of exposure limits. This research also helps to understand the phenotype of asthma caused by wood dust which is an area requiring further exploration.

## Introduction

Around 60,000 workers are employed in the UK woodworking industry generating an estimated annual turnover of around £4 billion (1). The Health and Occupational Research (THOR) surveillance scheme shows wood dust is a leading cause of occupational asthma (OA) in Great Britain. Exposure to wood dust is associated with an increased risk of asthma symptoms and abnormal physiology, in particular cough and bronchitis (2), airway hyperresponsiveness (3), excess lung function decline (4), as well as an increased risk of asthma (5). Respiratory symptoms, specific immunoglobulin E (SIgE), and OA have all been reported at wood dust exposures lower than the current UK workplace exposure limit (WEL) for softwood of 5mg/m^3^ and mixed/hardwood of 3mg/m^3^ (2). How the risk of OA changes across the spectrum of wood dust exposure is not well understood (6, 7), but recent observational follow-up supports a reduction in symptoms with reducing exposure (8).

Atopy has previously been shown to modify OA risk in woodworkers, and atopic workers with higher dust exposures have been shown to have a higher risk of asthma (3). Wood dust exposure appears to have a more significant impact on lung function among smokers, with the effect more pronounced at higher exposures (4). However, evidence supporting the influence of atopy and smoking on asthma risk in woodworkers is inconsistent and not reported in all studies (2, 9, 10).

Few recent studies have explored asthma risk in woodworkers, and no recent studies have been conducted in the UK (2). We aimed to identify risk factors for asthma in British woodworkers, with an emphasis on asthma symptoms, atopy and wood dust exposure.

## Methods

### 1.1 Study population

Worksites were identified through participation in a Health and Safety Executive (HSE) hygiene study and through the Corporate Operational Information System (COIN) database between 2014 and 2018 (1, 11). Worksites were representative of UK woodworking, ranging from small (<10 employees) to large (>200 employees) and from sectors including furniture manufacture, carpentry, boat building, and timber. All workers over 16 and currently exposed to any type of wood dust were eligible.

Full details of the hygiene study are published elsewhere (11). In brief, a passive sampling device was worn on the lapel for the duration of a working shift and standard eight-hour time-weighted averages (8-hr TWA) calculated for inhalable wood dust for the tasks sampled.

### 1.2 Health outcomes

Workers underwent an interviewer-administered questionnaire detailing symptoms based on the Medical Research Council (MRC) and European Community Respiratory Health Survey (ECRHS) questionnaires (12, 13).

‘Cough’ was defined as usual coughing during the day, ‘ever wheeze’ was defined as ever wheezing except during colds, ‘chronic bronchitis’ was defined as a cough with sputum for at least 3 months a year, ‘nasal symptoms’ were defined as reporting regular itchy, blocked or runny nose, and ‘ocular symptoms’ as regular symptoms of itchy, gritty, dry eyes, or conjunctivitis (8). Work-related nasal symptoms (WRNS), work-related ocular symptoms (WROS), and work-related respiratory symptoms (WRRS) were defined as worse at work or improving away from work or on holiday. Current asthma symptoms (CAS) were defined as wheezing, chest tightness, breathlessness, or asthma medication use within the last 12 months (13). Ever asthma was defined either current or past self-reported asthma diagnosis. Current asthma was defined as either a current or past physician-diagnosis of asthma along with CAS according to ERCHS criteria (13). Asthma with latency was defined as adult-onset asthma with onset after first exposure in the woodworking industry.

Fractional exhaled nitric oxide (FE_NO_) was performed before spirometry using a NOBreath device (Bedfont Scientific, Kent) according to American Thoracic Society/European Respiratory Society (ATS/ERS) standards (14). Spirometry was measured using an NDD Easy-On spirometer (Zurich, Switzerland) to ATS/ERS standards (15). Subjects were examined sitting and without a nose clip. Blood samples were analysed for total IgE (TIgE) and specific IgE to hard (oak, mahogany and obeche) and soft (beech, pine, cedar and silver fir) woods using standard ImmunoCAP testing (Phadia, Sweden, 2012). Forced expiratory volume in one second (FEV_1_), forced vital capacity (FVC), peak expiratory flow rate (PEF) and FEV_1_/FVC values falling two standard deviations below the mean (lower limit of normal or LLN) were considered abnormal (16). FE_NO_ values above 40ppb were considered high and values above 25ppb intermediate (17). Participants were considered sensitised if their IgE to hard or soft wood exceeded 0.35kU/L (18). Atopy was defined as a TIgE above 100 kU/L (19).

### 1.3 Statistical analysis

Wood dust exposures were assigned to each worker via a bespoke job exposure matrix (JEM) generated using a linear effects model, reported in full elsewhere (11). Not all worksites included in the hygiene study participated in the health study. Exposure was explored as continuous (geometric mean exposure) and categorical (quartiles of exposure, ‘high’ versus ‘low’ defined as the upper three quartiles versus the lowest quartile, and above or below a 2mg/m^3^, 3mg/m^3^ and 5mg/m^3^ exposure limit) variables (8).

Normally distributed data were displayed as means and standard deviations and compared using independent t-tests and ANOVA. Exposure, TIgE and FE_NO_ data were not normally distributed and reported as medians with interquartile range (IQR). Mann-Whitney-U tests were used to compare non-normally distributed data, and data were log transformed for use in regression models with back-transformed data reported in tables as geometric means. Categorical data were analysed using chi-squared tests.

Only technically acceptable spirometry and FE_NO_ measurements were used in the final analysis. The outcomes of interest including respiratory symptoms, WRRS, current or ever asthma, FE_NO_ >40ppb, and FEV_1_/FVC <LLN were used as dependent variables in logistic regression models. Wood dust exposure, atopy and CAS were identified a priori as explanatory variables increasing the risk of asthma in woodworkers. Models were adjusted for smoking, atopy, age, sex, and respiratory protective equipment (RPE) use (14). Stratified analyses were conducted to assess contribution of exposure, atopy and CAS on the dependent variables of interest (3). Crude and adjusted odds ratios (OR) were reported with associated 95% confidence intervals. All statistical analysis was performed using SPSS Statistics, v23 (20).

An NHS REC committee approved this research (reference 13/NI/0208).

## Results

Two-hundred-and-sixty-nine out of a possible 376 workers participated (participation rate 72%). Reasons for non-participation included inability to capture workers due to shift work, inability of workers to take time away from shift, worker annual leave, and worker refusal to participate.

Full results of the hygiene study are reported elsewhere (11). For worksites participating in the health study, median exposure to inhalable wood dust was 2.00 mg/m^3^ (IQR 1.14), and 18 (7%) of workers had a current exposure to wood dust above 3mg/m^3^. All workers used seasoned (dry) wood.

Table 1 shows key demographic, exposure, and clinical characteristics of the study population by presence of CAS. Most participants were male (n=261, 97%) with an average age of 42.4 (SD 12.6) years. Seventy (26%) workers were current smokers. One hundred and fifty four workers (57%) were atopic and one sixth of the study population (n=40, 15%) had current asthma. A significant proportion of the study population reported CAS (n=123, 46%). WRRS were less frequent at 11% (n=29). Thirteen workers (5%) had asthma with latency, with no difference between groups. Rates of SIgE sensitisation to wood dust were very low overall and not significantly different between groups.

**Table 1:** Key demographic, exposure, and clinical characteristics of 269 of British woodworkers, stratified by current asthma symptoms. Number of workers in each group is reported in parentheses.

|  | Current asthma symptoms<br>(n=123) | No current asthma<br>symptoms (n=146) | Total (n=269) |
| --- | --- | --- | --- |
| *Demographics* |  |  |  |
| Age, years (SD) | 42.3 (13.0) | 42.5 (12.2) | 42.4 (12.6) |
| Sex, m (%) | 118 (96) | 143 (98) | 261 (97) |
| Current smoker, n (%) | 39 (32) | 31 (21) | 70 (26)<sup>&</sup> |
| Ever smoker, n (%) | 39 (32) | 41 (28) | 80 (30) |
| *Exposure* |  |  |  |
| Hardwood exposure, n | 35 (28) | 41 (28) | 76 (28) |
| Softwood exposure n (%) | 14 (11) | 11 (8) | 25 (9) |
| Mixed exposure, n (%) | 74 (60) | 94 (64) | 168 (62) |
| Uses RPE, n (%) | 84 (68) | 123 (84) | 207 (77)<sup>**</sup> |
| Years exposed in woodworking<br>industry (SD) | 17.33 (12.50) | 20.21 (12.94) | 18.89 (12.80)<sup>*</sup> |
| Current exposure, $\text{mg}/\text{m}^3$ (IQR) | 2.0 (1.3) | 2.0 (0.9) | 2.0 (1.1) |
| Exposure $>5\text{mg}/\text{m}^3$, n (%) | 2 (1) | 1 (1) | 3 (1) |
| Exposure $>3\text{ mg}/\text{m}^3$, n (%) | 10 (8) | 8 (6) | 18 (7) |
| Exposure $>2\text{mg}/\text{m}^3$, n (%) | 65 (53) | 95 (65) | 160 (60)<sup>*</sup> |
| Exposure 'high' vs 'low', n (%) | 89 (72) | 113 (77) | 202 (75) |
| *Symptoms* |  |  |  |
| Ever wheeze, n (%) | 66 (54) | 18 (12) | 84 (31)*** |
| Cough, n (%) | 58 (47) | 24 (16) | 82 (31)*** |
| Chronic bronchitis, n (%) | 17 (14) | 4 (3) | 21 (8) |
| Nasal symptoms, n (%) | 36 (29) | 35 (24) | 71 (26) |
| Ocular symptoms, n (%) | 42 (34) | 25 (17) | 67 (25)*** |
| Any work-related respiratory symptom, n (%) | 27 (22) | 2 (1) | 29 (11)*** |
| Work-related nasal symptoms, n (%) | 16 (13) | 19 (13) | 35 (13) |
| Work-related ocular symptoms, n (%) | 34 (28) | 27 (19) | 61 (23)^ |
| **Asthma, atopy and sensitisation** |  |  |  |
| Current asthma, n (%) | 40 (33) | 0 (0) | 40 (15)*** |
| Ever asthma, n (%) | 40 (33) | 15 (10) | 55 (21)*** |
| Asthma with latency n (%) | 10 (8) | 3 (2) | 13 (5) |
| Atopic symptoms, n (%) | 67 (55) | 62 (43) | 129 (48) |
| Atopic, n (%) | 80 (65) | 74 (51) | 154 (57)* |
| Total IgE, Ku/L (IQR) n=246 | 46.0 (129.0) | 27.0 (62.0) | 36.0 (69.8)** |
| Positive SigE to hard or soft wood, n (%) | 0 (0) | 1 (1) | 1 (1) |
| **FE<sub>NO</sub> and spirometry** |  |  |  |
| FE<sub>NO</sub>, ppb (IQR)<sup>b</sup> | 19.3 (28.7) | 18.7 (19.8) | 18.7 (24.1) |
| FE<sub>NO</sub> >25ppb, n (%) | 47 (40) | 45 (32) | 92 (36) |
| FE<sub>NO</sub> >40ppb, n (%) | 27 (23) | 23 (17) | 50 (19) |
| FEV<sub>1</sub>, L (SD) | 3.7 (0.6) | 4.0 (0.7) | 99.8 (12.7)* |
| FEV<sub>1</sub> <LLN, n (%) | 2 (2) | 1 (1) | 3 (1) |
| FEV<sub>1</sub>/FVC <LLN, n (%) | 8 (7) | 4 (3) | 12 (4) |
| b n=258 |  |  |  |
| *p<0.05, **p<0.01 ***p<0.001, &p=0.051, ^p=0.074 |  |  |  |

Symptomatic workers had worked fewer years in the woodworking industry, were less likely to use RPE, and fewer were exposed at levels >2mg/m^3^. Wheeze, cough, ocular symptoms, and WRRS were more common among workers with CAS. Atopy and asthma were significantly more common in those with CAS: 33% of workers with CAS reported ever asthma versus 10% without (p<0.001).

No significant relationships between exposure and respiratory symptoms, WRRS or asthma were observed in regression models (table 2). In logistic regression models no positive dose-response was seen between increasing exposure to wood dust (either in quartiles of exposure, ‘high’ versus ‘low’, or above the 2mg/m^3^ threshold) and respiratory symptoms, CAS, asthma, airway inflammation or airflow obstruction (tables A1-A4, online appendix). Workers with exposures above 2mg/m^3^ were at significantly increased risk of WR nasal symptoms compared to those below the 2mg/m^3^ threshold (adjusted OR 3.08, 1.55 – 6.12, p<0.001, table A4, online appendix). Conversely, workers in the ‘high’ exposure group and workers with exposures above 2mg/m^3^ were at lower risk of respiratory symptoms, CAS, and WRRS compared to those in the lower exposure groups (tables A2 and A4, online appendix).

**Table 2:** Adjusted regression models for geometric mean wood dust exposure, respiratory symptoms, and asthma. Odds ratios and 95% confidence intervals are shown in parentheses.

|  | Ever wheeze | Chronic bronchitis | Nasal symptoms | Ocular symptoms | Any WRRS | WR nasal or ocular symptoms | Current asthma symptoms | Current asthma |
| --- | --- | --- | --- | --- | --- | --- | --- | --- |
| **GM Current exposure, mg/m<sup>3</sup> (95%CI)** | 1.63 (0.01 – 52.00) | 1.48 (0.03 – 77.98) | 0.54 (0.01 – 20.99) | 0.63 (0.01 – 77.27) | 0.79 (0.02 – 31.05) | 0.55 (0.01 – 74.82) | 0.27 (0.01 – 22.39) | 7.41 (0.08 – 693.43) |
| Models controlled for age, smoking, and RPE use. R<sup>2</sup> = 0.13 |  |  |  |  |  |  |  |  |

Atopic woodworkers were at significantly increased risk of nasal and ocular symptoms, WRRS, CAS and current asthma (table 3). However no specific associations were seen between atopy and cough, ever wheeze, or chronic bronchitis. Atopic woodworkers were at significantly increased risk of airway inflammation defined by a FeNO >40ppb (unadjusted OR 2.00, 1.03 – 3.88 p = 0.04), but no effect was seen for airflow obstruction (adjusted OR 0.8, 0.24 – 2.69).

**Table 3:**
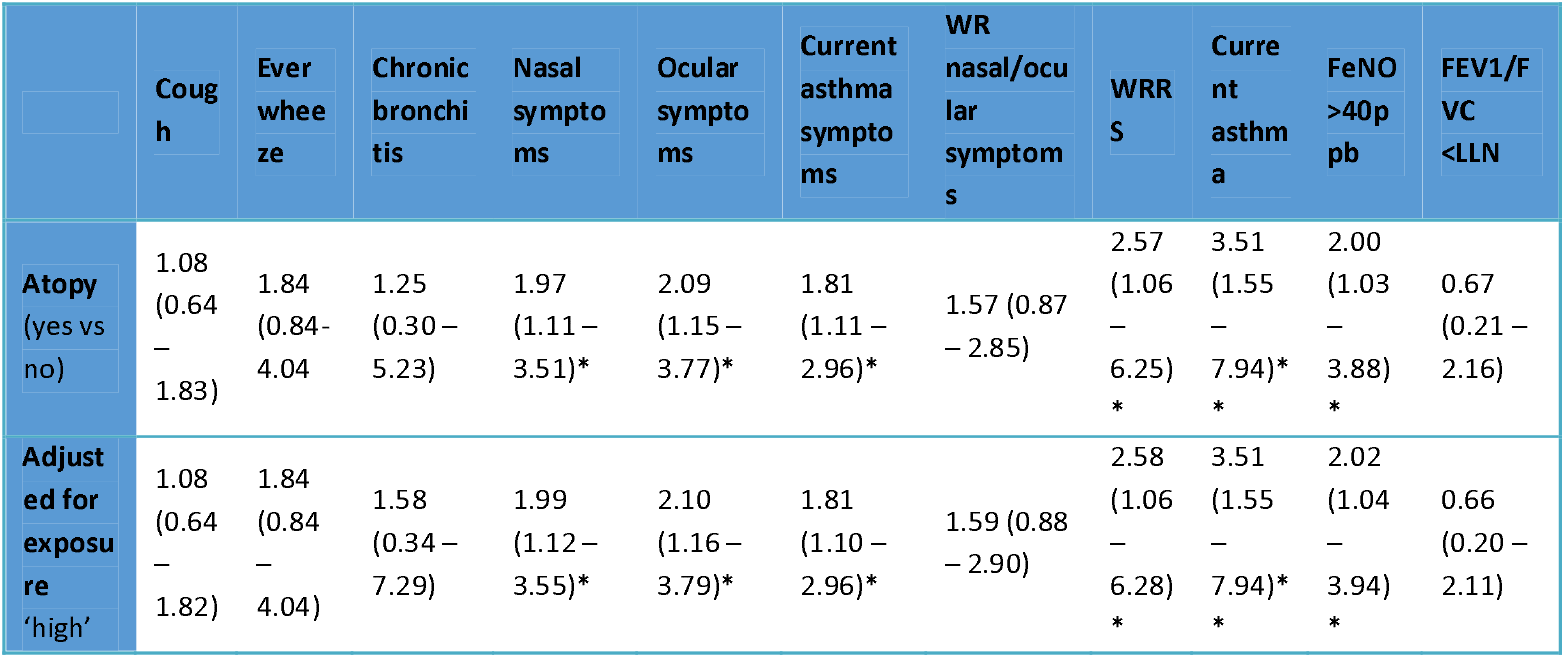
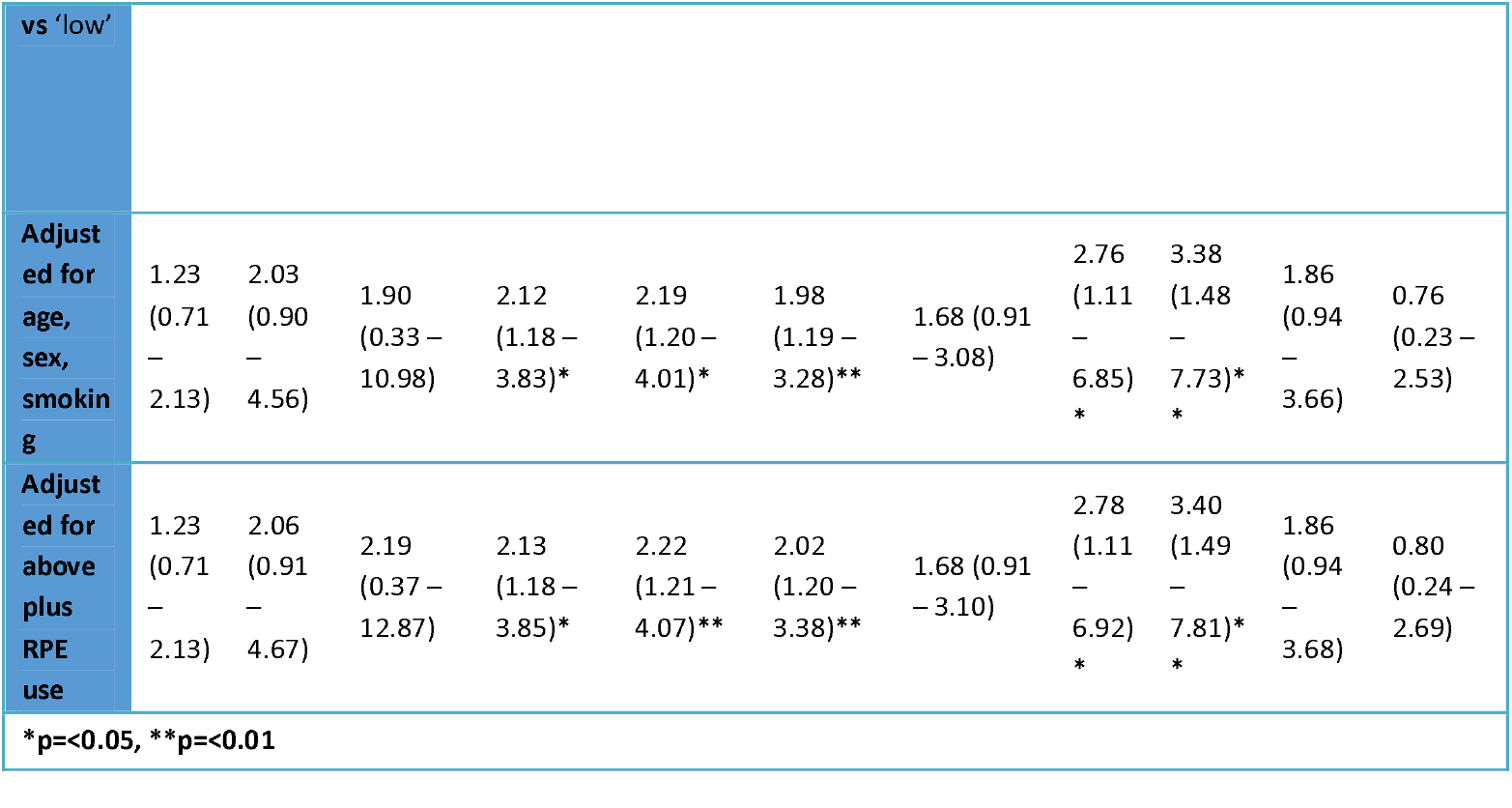
Crude and adjusted logistic regression models showing respiratory symptoms, work-related symptoms, airway inflammation and airflow obstruction in workers with atopy. Workers with no atopy were the control group. Odds ratios and 95% confidence intervals are shown in parentheses.

Woodworkers with CAS were at increased risk of ocular symptoms and asthma (table 4). In controlled analyses symptomatic workers were at increased risk of ocular symptoms (OR 2.94, 1.59 – 5.43, p<0.001) and tended towards increased risk of WR nasal and ocular symptoms although this did not reach significance. Woodworkers with CAS were at significantly increased risk of WRRS, though confidence intervals were wide (23.75, 5.35 – 105.33, p<0.001). Workers with CAS were significantly more likely to self-report ever asthma (4.29, 2.12 – 8.69, p<0.001). No clear associations were seen for CAS and FE_NO_ or airflow obstruction.

**Table 4:** Crude and adjusted logistic regression models between current asthma symptoms, work-related symptoms, airway inflammation and airflow obstruction. Workers with no current asthma symptoms were the control group. Odds ratios and 95% confidence intervals are shown in parentheses.

|  | Nasal symptoms | Ocular symptoms | WR nasal/ocular symptoms | WRRS | Ever asthma^ | FE<sub>NO</sub> >40ppb | FEV1/FVC <LLN |
| --- | --- | --- | --- | --- | --- | --- | --- |
| CAS (yes vs no) | 1.31 (0.76 – 2.26) | 2.51 (1.42 – 4.44)** | 1.68 (0.95 – 2.99) | 20.25 (4.71 – 87.14)*** | 4.18 (2.17 – 8.03)*** | 1.48 (0.80 – 2.75) | 2.60 (0.76 – 8.88) |
| **Adjusted for exposure** | 1.35 (0.78 – 2.33) | 2.57 (1.45 – 4.55)** | 1.77 (0.99 – 3.16) | 19.92 (4.62 – 85.87)*** | 4.19 (2.17 – 8.07)*** | 1.53 (0.82 – 2.85) | 2.54 (0.74 – 8.72) |
| **Adjusted for age, sex, smoking, BMI** | 1.36 (0.78 – 2.36) | 2.75 (1.53 – 4.94)*** | 1.77 (0.98 – 3.19) | 21.76 (5.00 – 94.57)*** | 4.84 (2.45 – 9.56)*** | 1.79 (0.94 – 3.40) | 2.04 (0.58 – 7.26) |
| **Adjusted for atopy** | 1.22 (0.69 – 2.15) | 2.56 (1.40 – 4.57)** | 1.66 (0.91 – 3.01) | 19.55 (4.48 – 85.24)*** | 4.31 (2.15 – 8.64)*** | 1.65 (0.86 – 3.18) | 2.22 (0.61 – 8.09) |
| **Adjusted for above plus RPE use** | 1.28 (0.72 – 2.30) | 2.94 (1.59 – 5.43)*** | 1.82 (0.99 – 3.36) | 23.75 (5.35 – 105.33)*** | 4.29 (2.12 – 8.69)*** | 1.82 (0.93 – 3.59) | 1.84 (0.49 – 6.93) |
| **p<0.01 ***p<0.001 |  |  |  |  |  |  |  |
| ^Ever asthma used as dependent variable due to model generation |  |  |  |  |  |  |  |

Stratified analyses were conducted to explore associations between symptoms and atopy and controlled for exposure and other confounders (table 5). Woodworkers with both CAS and atopy were more likely to have WRRS (OR 8.61, 3.55 – 20.84, p<0.001), current asthma (OR 15.86, 6.82 – 36.89, p<0.001), and a high FE_NO_ >40ppb (OR 2.44, 1.27 – 4.67, p<0.01). No associations were seen for asthma with latency or airflow obstruction. In workers with CAS and no atopy, no significant associations were seen with variables of interest. In the absence of CAS, atopic woodworkers were significantly less likely to have WRRS (0.08, 0.01 – 0.59, p<0.05) and no relationship was seen with current asthma or FE_NO_.

**Table 5:** Adjusted logistic regression models showing risk of work-related respiratory symptoms, asthma, airway inflammation and airflow obstruction stratified by presence of atopy and/or current asthma symptoms. Odds ratios and 95% confidence intervals are shown in parentheses.

|  | WRRS | Current asthma | Asthma with latency | FE<sub>NO</sub> >40ppb | FEV1/FVC<LLN |
| --- | --- | --- | --- | --- | --- |
| **CAS and atopy** | 8.61 (3.55 – 20.84)*** | 15.86 (6.82 – 36.89)*** | 1.06 (0.29 – 3.80) | 2.44 (1.27 – 4.67)** | 1.72 (0.51 – 5.76) |
| **CAS no atopy** | 1.43 (0.52 – 3.88) | 1.36 (0.57 – 3.26) | 1.01 (0.14 – 7.12) | 0.52 (0.17 – 1.58) | 1.38 (0.37 – 5.81) |
| **Atopy no CAS** | 0.08 (0.01 – 0.59)* | NS | 1.16 (0.27 – 5.22) | 0.76 (0.36 – 1.58) | 0.22 (0.23 – 1.80) |
| *p<0.05, **p<0.01, ***p<0.001 & p=0.81<br>Models controlled for age, sex, smoking, RPE use and wood dust exposure ('high' vs 'low') |  |  |  |  |  |

## Discussion

Asthma symptoms were prevalent among British woodworkers, even at low exposure levels. Symptoms of cough and wheeze were present in around 30% of the study population, and asthma was present in 15%, levels higher than would be expected in adults in the general population (21) and in similar proportion to those reported in a recent study of Scandinavian woodworkers (8). Atopy was a significant modifying factor in the risk of asthma symptoms, WRRS, asthma and airway inflammation. When stratified by the presence or absence of atopy, woodworkers with CAS were at significantly increased risk of WRRS, current asthma and airway inflammation only in the presence of atopy.

Asthma symptoms were prevalent in the current study, with rates comparable to recent systematic reviews (2, 9). Woodworkers have been demonstrated to be at increased risk of asthma symptoms in several workplace studies (5, 8, 22, 23). Female, but not male, woodworkers with higher exposures have been shown to have an increased risk for chronic bronchitis, though data from other studies is conflicting (24). Since female representation in the current study was very low (3%), analysis on the effects of gender was not possible. It is important for individuals, employers, occupational health practitioners and respiratory physicians to be aware of the risk of asthma in woodworkers of both genders and consider measures that may reduce this risk.

Atopic woodworkers were at increased risk of nasal and ocular symptoms, WRRS, CAS, and asthma. In analyses stratified by atopy and CAS, symptomatic and atopic woodworkers were significantly more likely to have WRRS suggesting that these individuals may be at increased risk of occupational asthma. Similarly, the same group of workers were at significantly increased risk of asthma. This is in contrast to previous research suggesting non-atopic (rather than atopic) woodworkers are at increased risk of WRRS (3). The current study differs through a smaller proportion of female workers (3% in the current study versus 26%), wood species exposure (mixed vs solely soft wood), and dust exposure levels (median 2mg/m^3^ vs 0.96 mg/m^3^). Higher dust exposures or mixed species exposure (in particular to wood composites) may explain these differences in findings. Atopy is well known to be a risk factor for non-occupational asthma, in particular ‘Th-2high’ disease. Since pre-employment screening based on atopy is unlikely to offer advantages to individual workers, pre-selection of workers away from woodworking based on atopic status is not recommended (25).

Wood dust contains both high-molecular weight (HMW) and low-molecular weight (LMW) allergens (26). A minority of wood species are associated with immunological proteins and production of SIgE that is more often seen in HMW OA (2). In contrast, mechanisms in western red cedar OA are not associated with SIgE and associated with plicatic acid (12). Wood dust has been associated with early, late, and dual specific inhalation challenge (SIC) responses (27, 28). Atopy is associated with an increased risk of OA particularly in HMW but not LMW-exposed workers (29). The low levels of specific sensitisation observed in the current study support the lack of a measurable IgE response.

Atopic workers were also at an increased risk of significant eosinophilic airway inflammation defined by a FE_NO_ >40ppb. Atopic individuals are well-documented to have a higher FE_NO_ (14). Airway inflammation was common among the study population and almost a fifth of workers had a FE_NO_ s>40ppb. A FE_NO_ >40 ppb has been shown to have a sensitivity of 78.6 - 88.3% and specificity of 82.6 - 89.5% for predicting asthma in symptomatic individuals (21). In the current study 23% of workers with CAS had a FE_NO_ >40ppb and 40% had a FE_NO_ >25ppb. Workers with CAS and atopy were significantly more likely to have a FE_NO_ >40ppb in adjusted analyses (OR 2.44, 1.27 – 4.67, p<0.01). In contrast, no significant relationships were observed with airflow obstruction. Spirometry is more commonly used as both a diagnostic and surveillance tool in populations at risk of OA. FE_NO_ has been demonstrated to identify early bronchial hyperresponsiveness in groups of occupationally exposed apprentices, even when spirometry is normal (30), and it is now recommended as a first line objective test for asthma in recently updated UK guidance (31). The development of obstructive lung function is a late sign in OA and, when present, may be irreversible (32). The utility of FE_NO_ in occupational settings is not established, but this research suggests it could be useful in the identification of early airway inflammation and should be further explored.

Most participants had neither a raised FE_NO_ nor obstructive airways disease. Although these measurements were made cross-sectionally, the absence of airway inflammation or airflow obstruction may suggest an explanation other than airways disease for respiratory symptoms (10). Respiratory irritants, alternative respiratory diagnoses, or alternative exposures may account for the presence of symptoms and longitudinal studies are needed to examine whether workers will develop lung disease (3).

Exposure to wood dust, either as a continuous or dichotomised variable, was not clearly associated with an increased risk of asthma symptoms or asthma but was associated with an increased risk of WR nasal and ocular symptoms and a reduced risk of WRRS. In the current study workers were also exposed to other irritants or allergens including glues, solvents, paints, and resins, and a small number also worked with isocyanates. Exposure attribution may therefore explain the lack of dose-response. Current dust exposures were used in analyses, but no data was available on previous dust exposures. The intensity of exposure to wood dust may be as important as overall exposure when assessing asthma risk and longitudinal exposures would enhance future exposure attribution and assessment (33).

The lack of a clear dose-response relationship may also be explained by the healthy worker effect (HWE) (34). Workers with CAS had significantly less time exposed in the woodworking industry suggesting attrition of workers with respiratory symptoms. Furthermore, there were few people within the study with airflow obstruction, again suggesting these people have left the workplace. Although OA can occur after many years exposure to an occupational allergen, it is more common in the first years of exposure (35). Workers had spent an average of 19 years within the woodworking industry, increasing the influence of the HWE. Cohort studies are required to further examine the HWE phenomenon among woodworkers, especially at lower exposure levels.

Requirements under the Control of Substances Hazardous to Health (COSHH) 2002 regulations require exposure to occupational asthmagens to be as low as reasonably practicable (ALARP). We found median wood dust exposures of 2.0 mg/m^3^, lower than both the 3mg/m^3^ mixed/hardwood and 5mg/m^3^ softwood UK WELs (11, 36). Our findings support data from the UK and across Europe suggesting there has been a reduction in wood dust exposure over time, reflecting changes in processing, employment, and safety measures (37). Data suggest the UK has higher average wood dust exposures compared to other European countries, which may explain previously observed high levels of wood dust OA (37).

### Limitations

Cross-sectional studies of this kind are liable to exposure misclassification and are influenced by other factors such as workforce, factory size, and health and safety practices (38). Exposure misclassification on polytomous scales may dilute effect sizes at higher exposures, and thus lead to error biases towards the null (39). We used a detailed JEM based on empirical measurements to reduce risk of exposure misclassification and included health and safety practices in models to reduce confounding. Measures of airway inflammation and lung function were only collected at a single time point, compared to symptom data which spanned 12 months or more. Longitudinal studies are needed to explore whether symptoms are related to development of abnormal airways physiology and to better understand the mechanisms of wood dust OA.

Definitions for atopy and sensitisation vary across studies and may impact comparability. Atopy is commonly defined through the presence of skin-prick positivity which was not performed in this study. However, a TIgE >100 kU/L has been shown to have good positive predictive value for at least one positive allergy test in individuals with asthma symptoms (19). Compared to skin-prick tests, total IgE correlates better with the total allergic component of asthma and is more easily comparable between populations (40). Since rates of SIgE sensitisation were low in the study it is unlikely they interfered with TIgE results. Even where specific wood allergens have been used in patients with confirmed wood dust OA, specific sensitisation rates have been low suggesting an IgE response is not significant (18). Where positive SIgE results are present they may be helpful in supporting a diagnosis of wood dust OA, but the current findings suggest that a negative IgE to wood dust is unlikely to be helpful in excluding wood dust OA.

## Conclusion

Current asthma symptoms are prevalent at levels of wood dust exposure lower than the current UK WEL. Atopy was a significant modifier for risk of asthma and airway inflammation among woodworkers, particularly among symptomatic workers. Further studies to phenotype and endotype populations of workers at risk of, and suffering from, OA will inform future approaches to screening and diagnosis in these populations. Longitudinal studies are required to monitor wood dust exposures on a local, regional, national and international scale to understand how this influences respiratory symptoms and disease in exposed populations.

## Data Availability

All data produced in the present study are available upon reasonable request to the authors.

## Notes

### Competing Interest Statement

The authors have declared no competing interest.

### Funding Statement

This study was funded by the Health and Safety Executive

### Author Declarations

Ethics committee/IRB NHS REC committee reference number 13/NI/0208 gave ethical approval for this work

