## Supplementary material for "Atopy, asthma symptoms, and eosinophilic airway inflammation in British woodworkers"

### APPENDIX

**Table A1: Work-related respiratory symptoms, atopy, current asthma symptoms, and asthma risk across different quartiles of wood dust exposure.** Odds ratios are displayed with 95% confidence intervals in parentheses.

| | Any WRRS | WR nasal symptoms | WR ocular symptoms | Current asthma symptoms | Self-reported asthma | Current asthma |
|---|---|---|---|---|---|---|
| Low vs lowest exposure | 0.39 (0.12-1.22) | 1.19 (0.34-4.17) | 1.34 (0.43-4.17) | 0.79 (0.37-1.68) | 1.90 (0.57-6.37) | 1.88 (0.69-5.11) |
| High vs lowest exposure | 0.45 (0.15-1.38) | 2.67 (0.86-8.35) | 0.39 (0.09-1.73) | 0.85 (0.39-1.86) | 0.78 (0.19-3.18) | 0.75 (0.24-2.33) |
| Highest vs lowest exposure | 0.16 (0.03-0.81)* | 2.44 (0.77-7.79) | 2.15 (0.68-6.84) | 0.83 (0.38-1.82) | 0.38 (0.07-2.19) | 0.55 (0.16-1.93) |
| Atopy yes vs no | 3.31 (1.29-8.48)* | 0.87 (0.33-2.29) | 2.73 (1.12-6.64)* | 2.07 (1.07-4.03)* | 3.95 (1.45-10.79)** | 4.30 (1.87-9.88)** |
| Ever smoker yes vs no | 0.34 (0.13-0.87)* | 0.77 (0.36-1.68) | 1.45 (0.63-3.36) | 2.14 (1.22-3.75)** | 0.65 (0.24-1.78) | 0.92 (0.40-2.07) |

*p<0.05; **p<0.01

Models also adjusted for age, BMI, inhaled steroid and RPE use, only significant predictors shown.

**Table A2: Crude and adjusted logistic regression models showing relationships between respiratory symptoms, work-related symptoms, asthma symptoms and asthma and exposure to wood dust. 'High' vs 'low' exposure.** Odds ratios are displayed with 95% confidence intervals in parentheses.

| | Ever wheeze | Chronic bronchitis | Nasal symptoms | Ocular symptoms | Any WRRS | WR nasal or ocular symptoms | Current asthma symptoms | Current asthma |
|---|---|---|---|---|---|---|---|---|
| Crude OR | 1.86 (0.90 – 3.83) | 0.60 (0.27 – 1.34) | 1.40 (0.66 – 2.99) | 1.11 (0.50 – 2.48) | 0.42 (0.17 – 1.06) | 2.12 (0.84 – 5.37) | 0.70 (0.33 -1.45) | 0.99 (0.41 – 2.39) |
| Adjusted for age, smoking | 1.58 (0.72 – 3.49) | 0.56 (0.24 – 1.30) | 2.10 (0.88 – 4.98) | 1.24 (0.51 – 3.00) | 0.36 (0.13 – 0.95)* | 1.35 (0.50 – 3.68) | 0.73 (0.32 – 1.66) | 1.21 (0.46 – 3.15) |

| | | | | | | | | |
|---|---|---|---|---|---|---|---|---|
| **Adjusted for RPE use** | 1.86 (0.89 – 3.89) | 0.66 (0.29 – 1.52) | 1.37 (0.64 – 2.93) | 1.04 (0.46 – 2.34) | 0.37 (0.14 – 0.94)** | 2.07 (0.81 – 5.24) | 0.80 (0.38 – 1.71) | 1.00 (0.41 – 2.42) |
| **Adjusted for atopy** | 1.51 (0.71 – 3.22) | 0.62 (0.27 – 1.41) | 1.54 (0.69 – 3.44) | 1.11 (0.48 – 2.57) | 0.38 (0.15 – 0.98)**** | 1.73 (0.67 – 4.47) | 0.71 (0.33 – 1.55) | 1.15 (0.44 – 2.98) |
| **Adjusted for all** | 1.66 (0.74 – 3.73) | 0.63 (0.27 – 1.48) | 2.07 (0.86 – 4.96) | 1.05 (0.42 – 2.60) | 0.29 (0.10 – 0.79)**** | 1.35 (0.49 – 3.67) | 0.87 (0.37 – 2.04) | 1.16 (0.43 – 3.08) |
| ***p = 0.038 **p=0.037 ***0.046 ****p=0.015** | | | | | | | | |

**Table A3: Crude and adjusted logistic regression models showing relationships for exposure to wood dust, airway inflammation and airflow obstruction. '**High' vs 'low' exposure. Odds ratios are displayed with 95% confidence intervals in parentheses**.**

| | **$FE_{NO}$ >25ppb** | **$FE_{NO}$ >40ppb** | **FEV1 <LLN** | **FEV/FVC <LLN** |
|---|---|---|---|---|
| **Crude OR** | 1.24 (0.52 – 2.99) | 1.40 (0.47 – 4.19) | 0.82 (0.07 – 9.68) | 0.64 (0.18 – 2.28) |
| **Adjusted for age, smoking** | 1.30 (0.54 – 3.14) | 1.46 (0.49 – 4.37) | 0.81 (0.70 – 9.59) | 0.57 (0.16 – 2.10) |
| **Adjusted for age, smoking, RPE use** | 1.24 (0.51 – 3.03) | 1.36 (0.45 – 4.11) | 0.80 (0.07 – 9.50) | 0.68 (0.18 – 2.56) |
| **Adjusted for age, smoking, RPE use, atopy** | 1.51 (0.59 – 3.88) | 1.33 (0.41 – 4.32) | 0.91 (0.08 – 11.20) | 0.62 (0.15 – 2.48) |
| **Adjusted for age, smoking, RPE use, atopy current asthma** | 1.50 (0.58 – 3.86) | 1.37 (0.42 – 4.46) | 0.94 (0.08 – 11.59) | 0.63 (0.16 – 2.54) |

**Table A4: Crude and adjusted logistic regression models showing relationships between respiratory symptoms, work-related symptoms, asthma symptoms and asthma and exposure to wood dust above or below the 2mg/m³ threshold.** Odds ratios and associated 95% confidence intervals shown.

| | **Ever wheeze** | **Chronic bronchitis** | **Chronic cough** | **Nasal symptoms** | **Ocular symptoms** | **Any WRRS** | **WR nasal or ocular symptoms** | **Current asthma symptoms** | **Current asthma** |
|---|---|---|---|---|---|---|---|---|---|
| **Crude OR** | 0.45 (0.22 – 0.95) | 0.35 (0.09 – 1.40) | 0.61 (0.36 – 1.04) | 1.60 (0.91 – 2.84) | 1.42 (0.80 – 2.52) | 0.44 (0.20 – 0.96)* | 3.17 (1.62 – 6.19)** | 0.60 (0.37 – 0.98)* | 0.64 (0.32 – 1.25) |
| **Adjusted for age, smoking** | 0.43 (0.20 – 0.91) | 0.28 (0.05 – 1.42) | 0.58 (0.34 – 1.00)^ | 1.61 (0.91 – 2.87) | 1.44 (0.80 – 2.57) | 0.44 (0.20 – 0.96)* | 3.18 (1.62 – 6.24)** | 0.59 (0.36 – 0.96)* | 0.63 (0.32 – 1.25) |

| | | | | | | | | | |
|---|---|---|---|---|---|---|---|---|---|
| **Adjusted for RPE use** | 0.49 (0.23 – 1.07) | 0.27 (0.04 – 1.90) | 0.60 (0.35 – 1.05) | 1.55 (0.87 – 2.78) | 1.31 (0.73 – 2.39) | 0.39 (0.18 – 0.88)* | 3.01 (1.52 – 5.94)** | 0.66 (0.39 – 1.09) | 0.68 (0.34 – 1.36) |
| **Adjusted for above plus atopy** | 0.49 (0.23 – 1.08) | 0.16 (0.01 – 1.79) | 0.61 (0.35 – 1.05) | 1.59 (0.88 – 2.88) | 1.34 (0.74 – 2.44) | 0.39 (0.17 – 0.89)* | 3.08 (1.55 – 6.12)** | 0.66 (0.39 – 1.10) | 0.69 (0.34 – 1.39) |
| | **^p=0.05 *p<0.05 **p<0.001** | | | | | | | | |
